# Forecasting off-target drug toxicity using proteomic and genetic data: insights from Torcetrapib

**DOI:** 10.64898/2025.12.03.25341213

**Authors:** Jenifer A. Brody, Colleen M. Sitlani, Bruce M. Psaty, Ting Ye, Peter Ganz, James S. Floyd, Neil M. Davies

## Abstract

In the development of new drugs, one of the leading causes of late-stage failures are off-target adverse effects, but they are difficult to predict before expensive large-scale clinical trials. Proteomic changes observed in randomized controlled trials (RCTs) and Mendelian randomization estimates of the effects of these changes can provide valuable evidence about the likely effects of drugs on health outcomes. We provide proof of principle for this approach using data from the ILLUMINATE trial of torcetrapib, a drug developed to increase high-density lipoprotein (HDL) cholesterol while reducing low-density (LDL) cholesterol, but which unexpectedly increased blood pressure and mortality. We used Mendelian randomization to estimate the causal effects of 95 proteins perturbed by 3 months of torcetrapib exposure on 19 health outcomes. Six proteins showed concordant effects with the results of the trial, including C-type mannose receptor 2 (MRC2), cGMP-specific 3’;5’-cyclic phosphodiesterase (PDE5A), Spondin-1 (SPON1), and Tyrosine-protein kinase receptor Tie-1 (TIE1), which increased blood pressure in the same direction as their observed protein. Our results demonstrate a generalizable genetic-proteomic framework for predicting likely adverse drug effects, reducing potential harm to patients and drug failure costs.

## Introduction

Serious, unanticipated off-target adverse drug effects are one of the key reasons for the late-stage failure of new pharmaceuticals.^1^ These costly failures can be detected during the conduct or after the completion of a Phase 3 trial or even after regulatory approval, once large numbers of patients have been exposed.^2^ High failure rates suggest that preclinical evidence from *in silico, in vitro* or animal studies used to inform drug development has limitations and can be an unreliable guide to efficacy and safety in humans.^3^ Although drug targets with supportive genetic evidence are more than twice as likely to succeed in regulatory approval,^4^ little is known about whether genetic approaches can inform potential off-target safety concerns. Large-scale proteomic profiling of blood samples before and after the intervention can identify key proteins that change in response to a drug and provide early insights into potentially concerning signals. Here, we aim to use Mendelian Randomization to estimate the potential causal effects of these proteins on long-term health outcomes, while identifying molecular mechanisms that lead to adverse outcomes.

Mendelian randomization, using genetic variation in the population as a natural experiment to estimate the causal effects of exposures,^5^ can help provide evidence to inform drug development.^6^ This approach commonly uses genome-wide association study (GWAS) summary data to estimate the effects of risk factors in the population, such as those of lipids, proteins, or other risk factors, on disease.^7^ Mendelian randomization can also be used to estimate the effects of perturbing individual drug targets, typically proteins, in an approach known as drug target Mendelian Randomization.^6^ An advantage of this approach is that it is efficient, inexpensive and fast because it uses GWAS summary data that have already been collected and publicly shared. It does not require interventions or de novo studies. Drug target Mendelian randomization has several limitations. First, it can only identify the lifetime effects of perturbing a target from conception, which may not reflect the effects of perturbing a target later in life.^8^ Second, there may be insufficient proteomic data on key targets of interest. Third, drug target Mendelian randomization estimates reflect the effect of specific drug targets, but not necessarily their aggregate effects across multiple targets and different tissues. Finally, all the targets affected by a given drug may not be well understood, or even known, skewing the information to fewer well-understood targets.

Proteomic array data are also increasingly used in pharmaceutical drug development and randomized controlled trials.^9,10^ Current panels can assay thousands of proteins, most commonly from blood samples.^11^ These panels can be used to assess differential levels of proteins in blood before and after a therapeutic intervention, in analyses that are adequately powered even in relatively small samples, given that each participant contributes data (unlike with clinical outcomes), readily providing evidence of the molecular pathways perturbed by a given drug. However, whether randomized or not, short-term proteomics data from pharmaceutical trials are not necessarily informative about the long-term health effects of perturbing a target. Inferring adverse effects from proteomic perturbations may be even more challenging, because in most cases it is unclear whether a given protein is a cause or a consequence of the off-target pathway.

Here, we propose leveraging proteomics from a clinical trial and Mendelian randomization to predict the long-term effects of perturbing the target with a pharmacological agent. This method can overcome potential limitations of the analysis of only the proteomic data from pharmaceutical trials, including concerns about causation, limited clinical follow-up, rare adverse effects, and inability to differentiate the target effects of the drug from the off-target effects of the drug molecule. It may also overcome some of the limitations of drug target Mendelian randomization using population-based data, such as a lack of specificity and knowledge of targets perturbed by a drug (**Figure 1**). By integrating trial data and Mendelian randomization, we aim to provide a fast, inexpensive, efficient, and statistically powerful method, especially for detecting rare clinical events.

**Figure 1:**
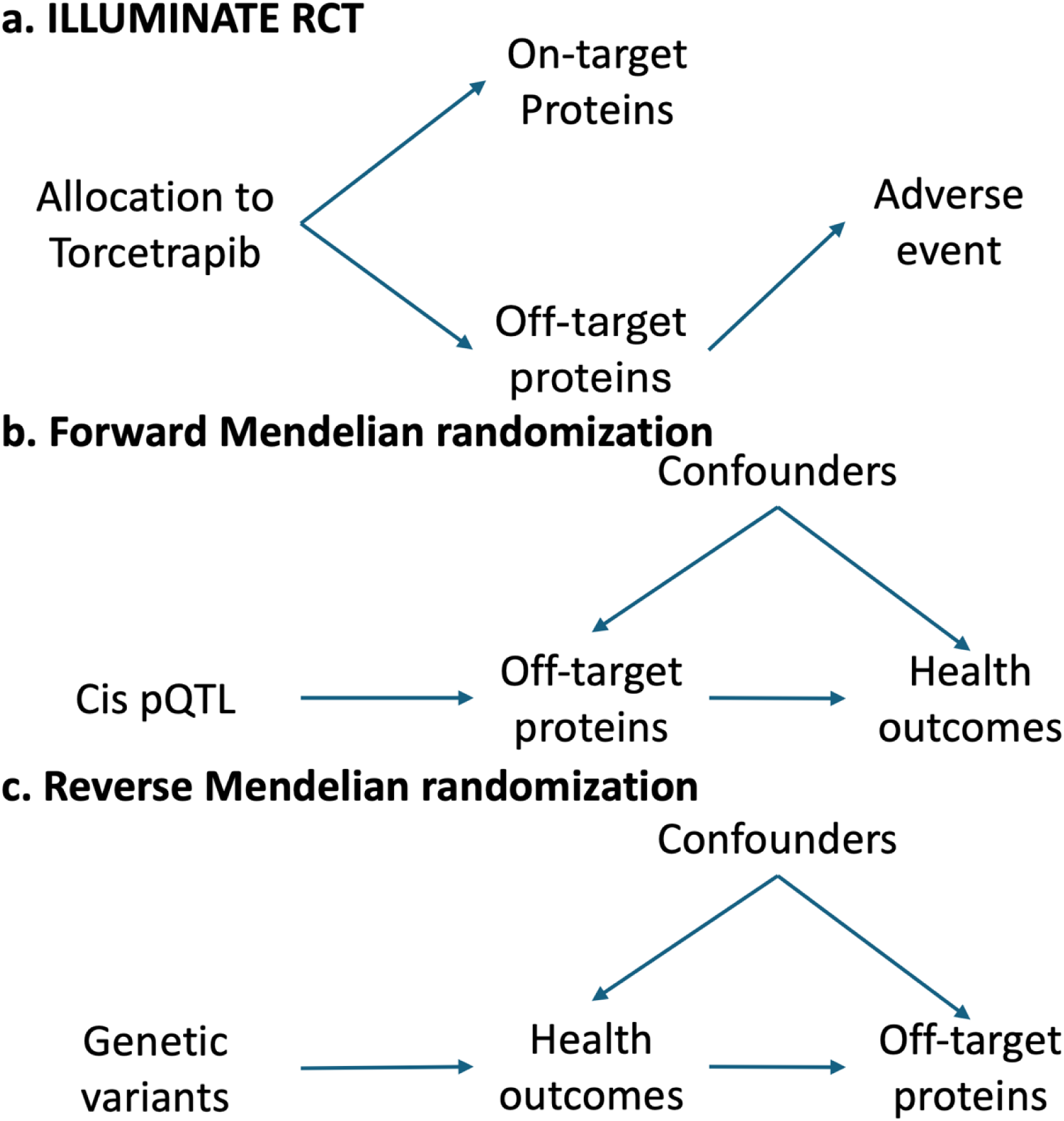
Directed Acyclic Graphs of the relationships between torcetrapib, the proteome, and cardiovascular outcomes. ILLUMINATE estimated the change in proteins after treatment with torcetrapib. Mendelian randomization estimates the life-long effects of genetic variants that perturb these proteins on health outcomes.

**Figure 2:**
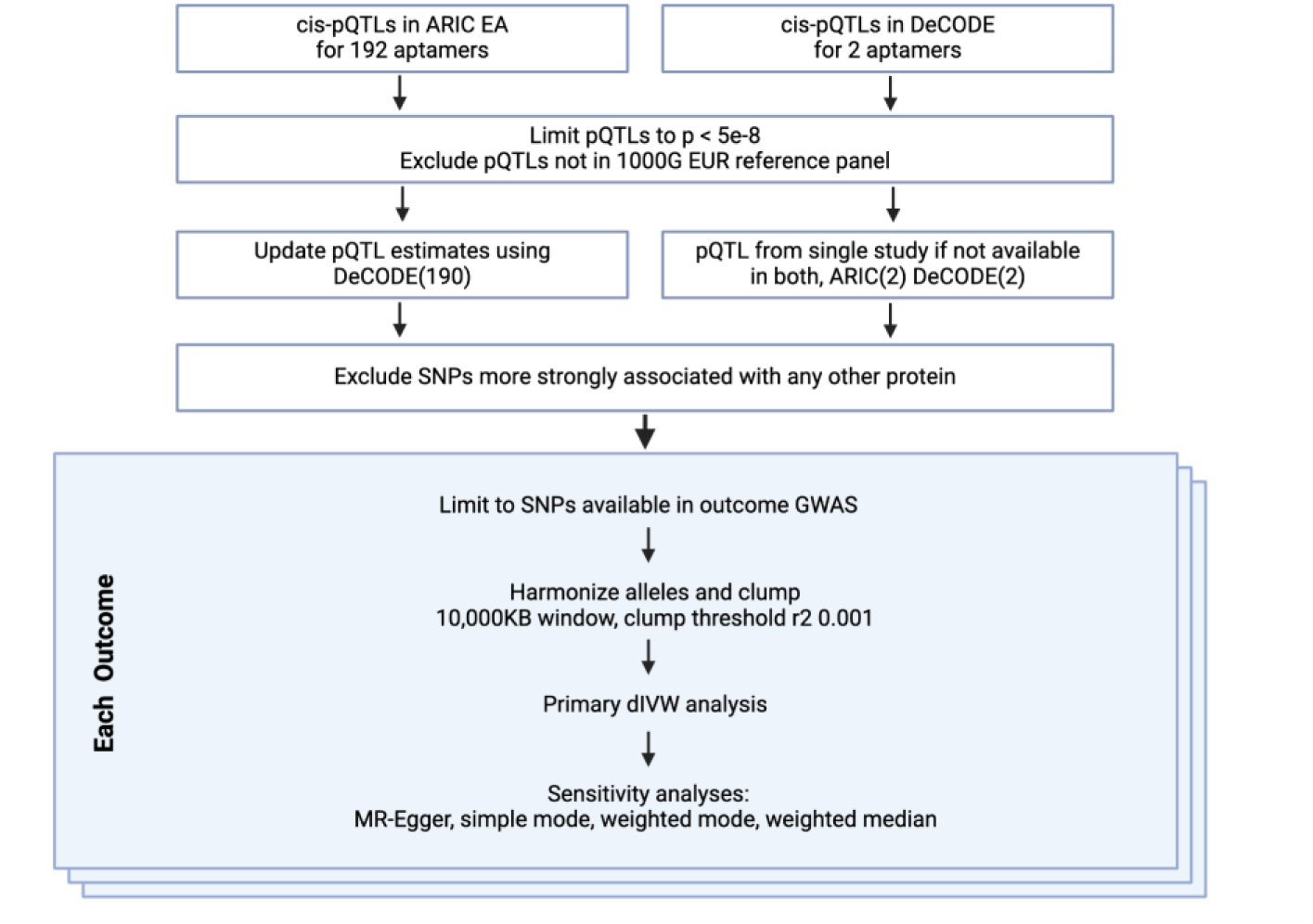
MR-STROBE Flow diagram of inclusion and exclusion of variants.

We illustrate this approach using torcetrapib, a cholesterol ester transfer protein (CETP) inhibitor, which aimed to prevent atherosclerotic cardiovascular disease by increasing HDL cholesterol while reducing LDL cholesterol, changes which are phenotypically associated with lower atherosclerotic cardiovascular disease rates. The phase three ILLUMINATE clinical trial randomized 15,067 participants to either torcetrapib and atorvastatin or atorvastatin alone.^12^ The trial was terminated early, because of a 25% (95%CI: 9% to 44%) increase in atherosclerotic cardiovascular events and a 58% (95%CI 14% to 139%) increase in all-cause mortality in those allocated to receive torcetrapib, as well as a 5.4 mmHg higher systolic blood pressure. A previous study used an early version of the SomaScan proteomics platform to measure 1,129 plasma proteins before and 3 months after treatment (n=249 participants in the active treatment arm), finding that 200 plasma proteins increased or decreased after treatment.^13^ In this present analysis, we used results from large external GWAS of several phenotypes and Mendelian randomization to investigate the potential causal role of these 200 plasma proteins in mediating off-target effects, including increased blood pressure, atherosclerotic cardiovascular events and mortality.

## Results

Our analyses began with the 200 proteins identified by Williams, Ganz and colleagues as differentially expressed between baseline and 3-month measurements in the torcetrapib arm of the ILLUMINATE trial.^13^ We identified genetic variants for these proteins using two large SOMAscan *cis*-pQTL studies and identified 201 aptamers on these platforms that mapped to these 200 proteins (**Supplemental Table 1**).

The primary targets of torcetrapib were expected to increase HDL-C and decrease LDL-C and triglyceride levels.^14^ Since the adverse effects were specific to torcetrapib and are likely not a class effect of CETP inhibitors,^17^ we used Mendelian randomization to identify the expected on-target causal effects of lipids (LDL, HDL, and triglycerides) on the plasma proteome, which were excluded from further analysis (n=70) (**Supplemental Figure 1**, **Supplemental Table 2**). Among the remaining 131 “off target” proteins, we identified independent *cis*-pQTLs (p<5×10^-8^) for 95 proteins (see **Methods**).

We then used Mendelian randomization to estimate the effects of each protein on each outcome to assess which, if any, proteins mediated the potential adverse off-target effects of torcetrapib. Mendelian randomization estimates the causal effect of the change in the protein level observed in the trial on a SD change in the outcome (continuous traits) or on the relative odds of a trait (binary traits). In addition to the increase in mortality, cardiovascular events, and blood pressure, a higher rate of cancer and infections/sepsis were observed in the torcetrapib arm. Therefore, we identified the largest available GWAS of cardiovascular diseases (coronary heart disease, ischemic stroke, heart failure & peripheral artery disease); metabolic traits/risk factors (HDL, LDL, triglycerides, heart rate, systolic and diastolic blood pressure, type 2 diabetes, estimated glomerular filtration rate (eGFR), body mass index (BMI)); infection/auto-immune diseases (infection, pneumonia, sepsis and inflammatory bowel disease) and cancer (breast and prostate) (**Supplemental Table 3**). We then used Mendelian randomization to estimate the effects of each protein on these 19 health outcomes. Of the 95 proteins, 17 affected one or more outcomes after multiple testing correction (**Figure 3**, **Table 1**). Of these, seven proteins showed concordant effects with trial observations, meaning the Mendelian randomization estimates matched the direction of effect of the observed phenotypic changes in the trial.

**Figure 3.**
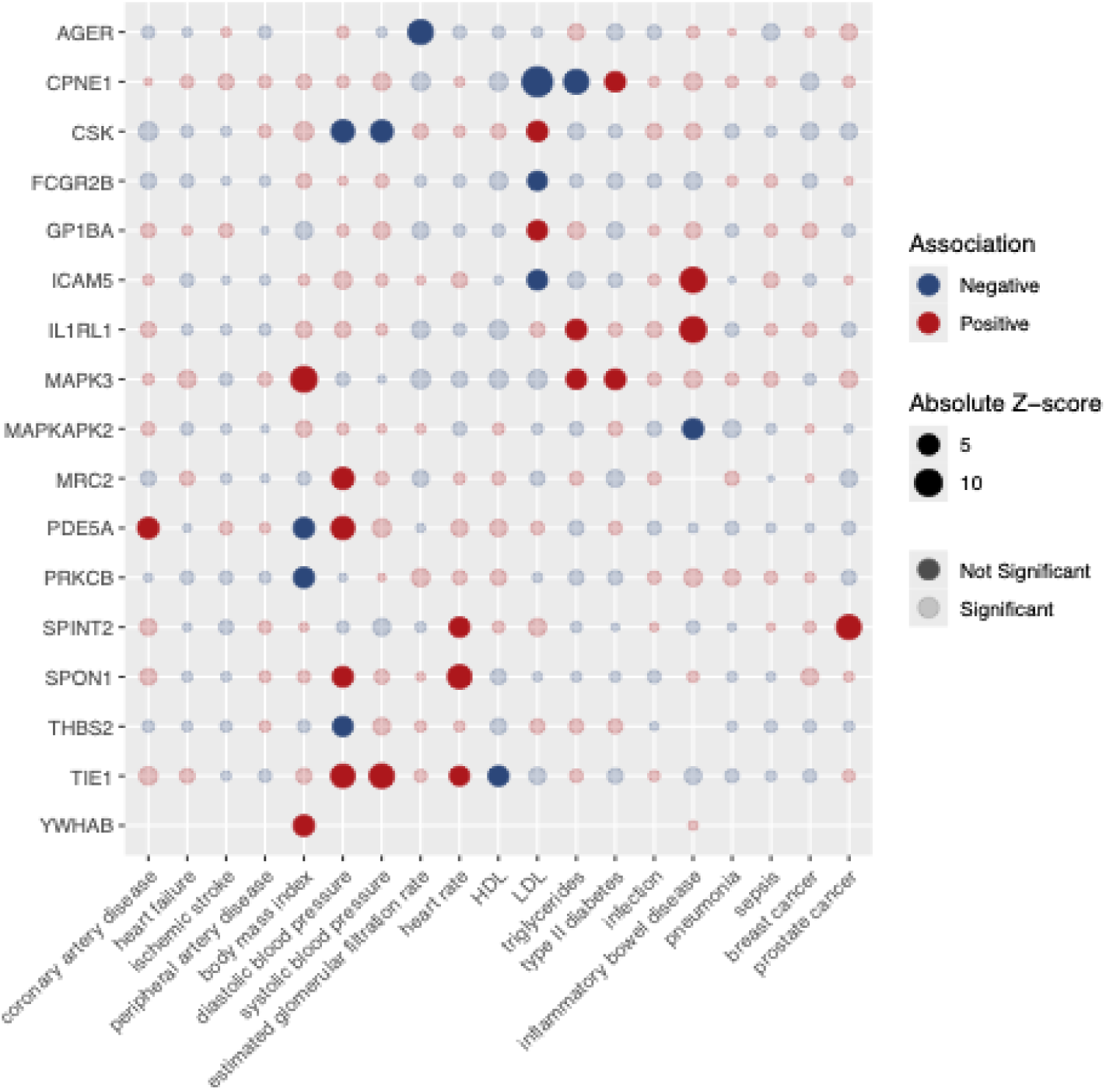
Mendelian randomization estimates of plasma proteins on health outcomes. Mendelian randomization estimates for 17 proteins showing significant effects on 12 of 19 different health outcomes after multiple testing correction (P<4.78E-5). Solid circles represent statistically significant associations after multiple testing correction; light-shaded circles represent non-significant estimates that were tested. Red circles indicate positive associations (genetically predicted protein changes in the same direction as observed in the trial associated with increased outcome risk/levels); blue circles indicate negative associations (genetically predicted protein changes in the same direction as observed in the trial associated with decreased outcome risk/levels). Effect sizes represent the change in outcome per standard deviation of the observed protein change in the trial, expressed per standard deviation change (continuous traits) or per odds ratio (binary traits).

**Table 1:** Concordance of the effects of torcetrapib-perturbed plasma proteins on cardiovascular, metabolic, immune, and cancer outcomes observed in the trial and Mendelian randomization.

| Protein exposure | SomaScan<br>SeqID | Outcome | N inst | F statistic | Observed<br>protein effect<br>(log2-fold change)<br>(1) | Trait change in<br>observational<br>trial (2) | Beta (3) | SE | P | Concordant<br>(4) | Outcome Type |
| --- | --- | --- | --- | --- | --- | --- | --- | --- | --- | --- | --- |
| AGER | 4125_52 | eGFRcrea | 1 | 1352 | 0.092 | Up | -0.029 | 0.004 | 4.41E-16 | 0 | continuous |
| CPNE1 | 5346_24 | LDL | 3 | 3026 | -0.262 | Down | -0.029 | 0.002 | 4.40E-42 | 1 | continuous |
| CPNE1 | 5346_24 | triglycerides | 3 | 3026 | -0.262 | Down | -0.017 | 0.002 | 1.53E-15 | 1 | continuous |
| CPNE1 | 5346_24 | type II diabetes | 3 | 3026 | -0.262 |  | 0.026 | 0.006 | 3.81E-06 |  | binary |
| CSK | 3363_31 | diastolic blood pressure | 1 | 53 | -0.246 | Up | -0.425 | 0.065 | 8.55E-11 | 0 | continuous |
| CSK | 3363_31 | LDL | 1 | 53 | -0.246 | Down | 0.159 | 0.033 | 1.27E-06 | 0 | continuous |
| CSK | 3363_31 | systolic blood pressure | 1 | 53 | -0.246 | Up | -0.306 | 0.05 | 1.20E-09 | 0 | continuous |
| FCGR2B | 3310_62 | LDL | 3 | 18092 | 0.044 | Down | -0.007 | 0.002 | 3.64E-05 | 1 | continuous |
| GP1BA | 4990_87 | LDL | 1 | 288 | -0.046 | Down | 0.047 | 0.011 | 9.88E-06 | 0 | continuous |
| ICAM5 | 5124_69 | inflammatory bowel disease | 1 | 1262 | 0.029 |  | 0.305 | 0.034 | 7.45E-19 |  | binary |
| ICAM5 | 5124_69 | LDL | 3 | 2426 | 0.029 | Down | -0.012 | 0.003 | 7.95E-06 | 1 | continuous |
| IL1RL1 | 4234_8 | inflammatory bowel disease | 1 | 18372 | 0.06 |  | 0.138 | 0.015 | 2.43E-20 |  | binary |
| IL1RL1 | 4234_8 | triglycerides | 2 | 9434 | 0.06 | Down | 0.01 | 0.002 | 1.10E-06 | 0 | continuous |
| MAPK3 | 2855_49 | body mass index | 1 | 822 | -0.223 |  | 0.078 | 0.008 | 1.31E-20 |  | continuous |
| MAPK3 | 2855_49 | triglycerides | 2 | 413 | -0.223 | Down | 0.028 | 0.006 | 8.52E-06 | 0 | continuous |
| MAPK3 | 2855_49 | type II diabetes | 2 | 413 | -0.223 |  | 0.081 | 0.017 | 1.03E-06 |  | binary |
| MAPKAPK2 | 3820_68 | inflammatory bowel disease | 1 | 376 | -0.187 |  | -0.313 | 0.064 | 1.00E-06 |  | binary |
| MRC2 | 3041_55 | diastolic blood pressure | 3 | 169 | 0.06 | Up | 0.057 | 0.01 | 4.73E-09 | 1 | continuous |
| PDE5A | 16805_5 | body mass index | 2 | 241 | -0.155 |  | -0.052 | 0.01 | 3.41E-07 |  | continuous |
| PDE5A | 16805_5 | coronary artery disease | 2 | 237 | -0.155 | Up | 0.15 | 0.029 | 1.60E-07 | 1 | binary |
| PDE5A | 16805_5 | diastolic blood pressure | 2 | 237 | -0.155 | Up | 0.064 | 0.01 | 1.56E-11 | 1 | continuous |
| PRKCB | 5475_10 | body mass index | 2 | 36 | -0.352 |  | -0.157 | 0.031 | 6.63E-07 |  | continuous |
| SPINT2 | 2843_13 | heart rate | 3 | 2412 | -0.052 |  | 0.017 | 0.004 | 4.61E-06 |  | continuous |
| SPINT2 | 2843_13 | prostate cancer | 2 | 3601 | -0.052 |  | 0.094 | 0.012 | 1.93E-15 |  | binary |
| SPON1 | 4297_62 | diastolic blood pressure | 2 | 638 | 0.063 | Up | 0.026 | 0.005 | 9.12E-07 | 1 | continuous |
| SPON1 | 4297_62 | heart rate | 2 | 638 | 0.063 |  | 0.049 | 0.007 | 2.59E-13 |  | continuous |
| THBS2 | 3339_33 | diastolic blood pressure | 4 | 1504 | 0.143 | Up | -0.016 | 0.004 | 8.75E-06 | 0 | continuous |
| TIE1 | 2844_53 | diastolic blood pressure | 1 | 575 | -0.054 | Up | 0.063 | 0.009 | 4.83E-13 | 1 | continuous |
| TIE1 | 2844_53 | HDL | 1 | 575 | -0.054 | Up | -0.036 | 0.008 | 2.33E-06 | 0 | continuous |
| TIE1 | 2844_53 | heart rate | 1 | 575 | -0.054 |  | 0.045 | 0.011 | 2.44E-05 |  | continuous |
| TIE1 | 2844_53 | systolic blood pressure | 1 | 575 | -0.054 | Up | 0.068 | 0.009 | 2.26E-15 | 1 | continuous |
| YWHAB | 14156_33 | body mass index | 2 | 195 | -0.168 |  | 0.057 | 0.011 | 2.16E-07 |  | continuous |

1) Log2-fold change in protein levels among participants between measures at baseline and three months in the torcetrapib arm
2) Change in trait among participants in the torcetrapib arm compared to the control arm of the ILLUMINATE trial
4) MR proteins are classified as concordant if the MR estimate is in the same direction as the trait change (column G) as observed in the trial

Six proteins had causal effects on systolic or diastolic blood pressure, four of which were concordant with the observed trial result of increased blood pressure: C-type mannose receptor 2 (MRC2), cGMP-specific 3’;5’-cyclic phosphodiesterase (PDE5A), Spondin-1 (SPON1), and Tyrosine-protein kinase receptor Tie-1 (TIE1) (all P<10⁻⁶). For example, MRC2 protein levels increased in response to torcetrapib (log2-fold change=0.06), and the Mendelian randomization estimates suggested that increased MRC2 levels increased diastolic blood pressure (β=0.057 SD units per SD increase in MRC2, P=4.73×10⁻⁹), concordant with the observed 2.1 mmHg diastolic blood pressure increase in the trial. Further, the observed decrease in PDE5A levels in the trial (log2-fold change=-0.155) is consistent with the Mendelian randomization effect of increased diastolic blood pressure (β=0.064 SD units per SD decrease in PDE5A, P=1.56×10⁻¹¹) per SD decrease in PDE5A levels. In contrast, Tyrosine-protein kinase CSK (CSK) and Thrombospondin-2 (THBS2) showed discordant effects with trial observations.

Mendelian randomization estimates for PDE5A showed trial-concordant effects on an increase in diastolic blood pressure. In addition to its effects on blood pressure, decreased PDE5A levels predicted increased coronary artery disease risk (β=0.15, P=1.60×10⁻⁷), consistent with the 25% increase in cardiovascular events observed in the trial. This finding appears paradoxical, given that PDE5A inhibitors such as sildenafil are vasodilators that lower blood pressure. However, our genetic instruments show context-dependent effects: the direction of association between genetically predicted PDE5A levels and outcomes varies by tissue and whether protein or transcript levels are being proxied (**Supplemental Table 4**).

We identified protein effects on metabolic outcomes that were not observed in the ILLUMINATE trial: Copine-1 (CPNE1) and Mitogen-activated protein kinase 3 (MAPK3) levels decreased after torcetrapib treatment, which increased type 2 diabetes risk (β=0.026, P=3.81×10⁻⁶ and β=0.081, P=1.03×10⁻⁶, respectively). Other proteins affected lipid levels, BMI, inflammatory bowel disease, prostate cancer risk, and kidney function. Complete statistical details, including effect sizes, confidence intervals, and instrument strength, are provided in **Table 1**.

Reverse Mendelian randomization was used to investigate the effects of each health outcome on the 131 “off-target” plasma proteins and, specifically, whether genetically-proxied mechanisms that lead to these outcomes are the cause of plasma protein perturbations from torcetrapib. These effects influenced 96 (73%) of the off-target proteins: cardiovascular diseases and metabolic traits explained 14 and 80 proteins respectively (89 combined across categories), while infection/autoimmune explained eight and cancer only one protein (**Figure 4** and **Supplemental Table 5**). Notably, some proteins were specific to certain health outcomes (e.g., coronary artery disease, diabetes), suggesting their potential as disease-specific surrogate endpoints. Most (79%) of the off-target protein changes observed in ILLUMINATE can be mechanistically linked to torcetrapib’s biological effects, either as mediators of adverse outcomes or as downstream markers of physiological changes.

**Figure 4.**
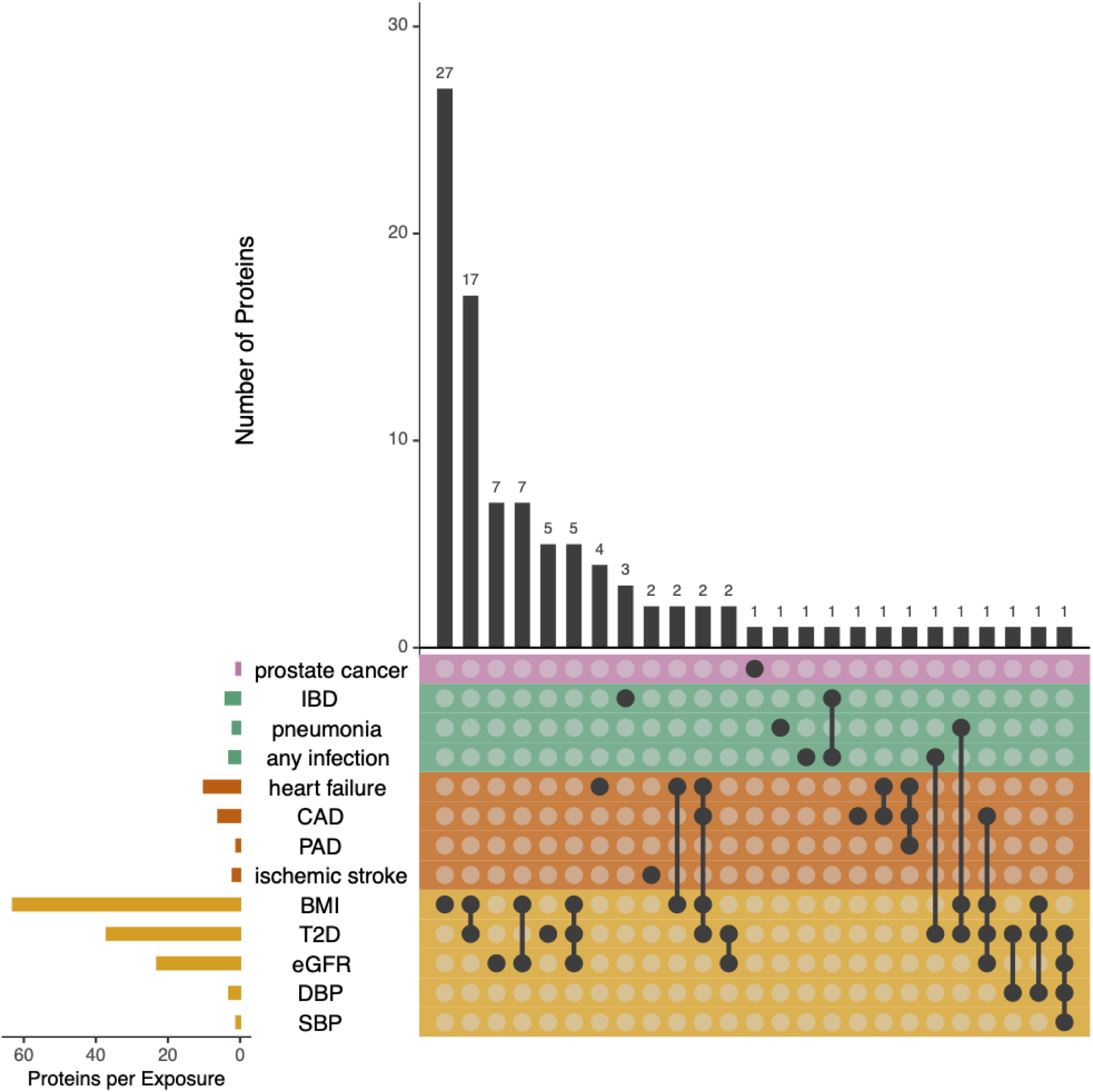
Reverse Mendelian randomization of health outcomes on protein levels. In the Upset plot each row represents a health outcome (cancer in pink, immune/infection in green, cardiovascular in orange, risk factors in yellow), and each column a protein with a valid genetic instrument. Filled black circles indicate significant Mendelian randomization estimates of the causal effect of genetically predicted disease or risk factors on plasma protein concentrations after Bonferroni correction; open circles indicate non-significant associations. The vertical bar plot (top) shows the number of proteins influenced by each outcome combination. In contrast, the horizontal bar plot (left) shows the number of proteins significantly affected by each outcome. Metabolic traits such as body-mass index (BMI), type 2 diabetes (T2D), estimated glomerular filtration rate (eGFR), systolic blood pressure (SBP), and diastolic blood pressure (DBP) exhibited the strongest and broadest proteomic influences.

For all analyses, we found little evidence that our conclusion differed when we used statistical approaches that made alternative assumptions about the structure of pleiotropy (**Supplemental Table 6**).

## Discussion

To the best of our knowledge, this is the first report to triangulate evidence from proteomic array data before and after a pharmaceutical intervention, with human genetics, and results from a large Phase 3 randomized controlled trial, in an effort to understand the biological mechanism leading to adverse drug effects. We identified 17 plasma proteins affected by torcetrapib in the ILLUMINATE trial that may be molecular mediators of the adverse health outcomes observed in the trial. A much larger number of protein changes appear to be the consequence rather than the cause of off-target effects from torcetrapib. These two distinct sets of protein biomarkers have different implications for early-stage drug development.

Biomarkers have long been used as surrogate outcomes to reduce sample sizes and shorten the duration of clinical trials before regulatory approval, and to identify promising new therapies in early-stage trials before long-term health outcomes are known.^15^ However, given strong assumptions about capturing intended and unintended causal effects, they come with the trade-off of uncertainty about the on- and off-target effects of these therapies, which may be inadequately understood before widespread use by patients. The first set of 17 plasma proteins we identified from Mendelian randomization effects of proteins on outcomes, which have strong empirical evidence of causal effects on adverse health outcomes from human genetics, can be used in small early-stage trials to select better therapies that are less likely to have harmful off-target effects to advance to costly Phase 3 trials, which are orders of magnitude larger and take years to complete. Treatments that decrease plasma levels of harmful proteins and increase levels of beneficial proteins may be more likely to be proven effective and safe in these trials than treatments with opposite effects on these Mendelian randomization-supported proteins.

The larger set of 96 proteins identified from reverse Mendelian randomization, although not causal effectors of adverse health outcomes, may be useful if they can reliably capture information about perturbations in biological mechanisms that lead to disease. The hundreds of variants that comprise our genetic instruments for health outcomes reflect heterogeneous and incompletely understood biological mechanisms. Similarly, our goal was to identify unique proteins or protein combinations sensitive to the various molecular and pathophysiological processes that can lead to target and off-target health outcomes. Notably, our approach leverages empirical evidence of interventional effects on the plasma proteome and therefore differs from widely used drug-target discovery approaches based solely on human genetics.^16^ Analyses limited to the intended molecular target of torcetrapib (CETP) have failed to recapitulate any of the drug’s adverse effects,^17^ while we have demonstrated that interrogating just a fraction of the human proteome in a small sample can identify dozens of potential molecular mediators or surrogate markers for important health outcomes.

### Strengths and Limitations

A strength of our study is the use of multiple complementary and sometimes contrasting forms of causal evidence, from proteomics and genetics, to identify several molecular mediators of well-defined adverse drug effects, as well as several promising biomarkers that merit further study as potential surrogate outcomes in cardiovascular clinical trials. We note several important limitations in this brief letter. This older version of the SomaScan platform measured only 1,129 proteins, compared with 11,000 with the latest generation, and all highly multiplexed affinity-based assays have the limitation of incomplete validation of the specificity of protein targets.^18,19^The proteins we investigated, which were affected by 3 months of torcetrapib treatment, represent either causes or consequences of this drug’s off-target effects. With reverse Mendelian randomization, we found that 9 of the 17 causal proteins showed little evidence of being affected by outcomes, supporting their role as upstream mediators, while 8 showed potential bidirectional relationships. Moreover, we identified several proteins that may represent attractive surrogate cardiometabolic outcomes. RCTs identify the short-term effect of a drug target using a specific molecule, which, for many drugs like torcetrapib, potentially affects numerous biological pathways and proteins in the blood. Meanwhile, Mendelian randomization estimates the lifetime effects of perturbing a single protein in the tissue measured in the proteomics GWAS.^20^ Thus, the ILLUMINATE and Mendelian randomization estimates may not reflect the same population parameter (estimand). Finally, all our proteomics data have come from blood plasma. If the adverse effects of torcetrapib are solely due to its effects in another tissue matrix, we may not have detected them.

### Future research

Future research should investigate whether proteomics data and Mendelian randomization can be used to forecast an intervention’s beneficial and adverse effects based on effects on the plasma proteomics in hundreds of patients, before thousands or tens of thousands are exposed in expensive Phase 3 trials. Further studies could use simulations to investigate the relative power of conventional RCTs versus proteomics-based approaches, as proteomic data may improve power and detection of beneficial or adverse effects in smaller samples.

## Conclusions

Proteomics data from RCTs can identify proteins affected by different treatments. This, when combined with Mendelian randomization can provide estimates of the likely long-term effects of perturbing a given drug target. This approach could improve the safety of new drugs and identify existing drugs that could be repurposed for new indications.

## Methods

We identified the proteins perturbed by torcetrapib using summary data from the ILLUMINATE randomized trial. We then estimated the effect of perturbing these proteins on cardiovascular outcomes: coronary heart disease (CHD), ischemic stroke (IS), heart failure (HF), peripheral artery disease (PAD), CHD risk factors, immune and cancer phenotypes. First, we identified all proteins affected by torcetrapib using SomaLogic proteomics data. We then estimated the effects of perturbing each protein on health outcomes using two-sample Mendelian randomization. We rescaled the Mendelian randomization estimates to reflect the size of the change in protein seen in the trial. Below, we describe the proteins, GWASs, genetic variants, and estimators used.

### Study design and data sources

#### ILLUMINATE RCT

ILLUMINATE randomized 15,067 participants to either torcetrapib and atorvastatin or atorvastatin alone between August 23, 2004, and December 28, 2005. The study recruited men and women between the ages of 45 and 75 with a history of cardiovascular disease or type 2 diabetes on hyperglycemic treatment in the 30 days to 5 years before eligibility screening. The original study design was planned for a follow-up period of approximately 4.5 years. Participants were excluded if they had an unstable medical condition, a short life expectancy, untreated LDL cholesterol levels of less than 2.6 mmol/L, or if they experienced a cardiovascular event during the 10-week run-in period to establish the atorvastatin dose needed to achieve LDL levels of less than 2.6 mmol/L. A paired subsample of 494 participants had blood samples taken at baseline and three months later. 247 individuals were selected as cases who experienced myocardial infarction, stroke/transient ischemic attack, hospitalization for heart failure, or all-cause death after the first three months, and these were matched to 247 controls who did not have an event at any point in follow-up. Controls were matched on atorvastatin dose after the 10-week run-in period, coronary heart disease, diabetes status, age, sex, and censoring time. The baseline blood sample was taken after the 10-week run-in period. Plasma (EDTA) was extracted and analyzed for 494 individuals, each with two blood samples, providing data on 1,129 proteins. After QC and removal, 986 proteins from 472 individuals were analyzed. Of these proteins, 200 were changed after treatment with torcetrapib. We identified the 200 proteins that changed after torcetrapib treatment and extracted their UniProt ID, log-2-fold change, and False-Discovery-Rate p-value.

#### Mendelian randomization

Mendelian randomization is an application of instrumental variable analysis that uses genetic variants as instrumental variables for the exposure of interest, here proteins^.21^ Instrumental variables are defined by three assumptions: 1) relevance, that the genetic variant associates with the exposure of interest, 2) independence, that there are no uncontrolled confounders or controlled colliders of the instrument-outcome association, and 3) exclusion: that all of the effects of the instrument are mediated via the exposure of interest. We describe how we assessed each of these assumptions below.

#### Proteomics GWAS

We wanted to focus on torcetrapib’s off-target effects, i.e., those not mediated via its effects on lipids. Therefore, we excluded proteins that were affected by lipid levels. For the remaining proteins unaffected by lipids, we identified independent *cis*-pQTLs in the Atherosclerosis Risk in Communities (ARIC) Study European-ancestry sub-sample (N=7,213).^<u>22</u>^ We used GWAS summary statistics for the proteomics data in ARIC for 4,483 proteins from blood plasma.^<u>22</u>^ We then selected genetic variants associated with each protein at genome-wide significant levels (p<5×10^-8^) and clumped to independent variants (r^2^<0.001) across 10,000kb using the TwoSampleMR package.^<u>23</u>^ To reduce the possibility of winner’s curse and to increase precision, we then identified the associations of each SNP with each protein using summary data from deCODE (N=35,559).^<u>24</u>^ While we cannot test whether the pQTLs have comparable effects in the outcome GWAS, pQTLs have been widely replicated across samples. To date, there is no evidence of substantial heterogeneity. Mendelian randomization estimates are reported as the effect of protein exposure on the outcome, expressed per standard deviation (SD) change in protein level in the direction of the observed protein effect. For proteins that increased after torcetrapib treatment, beta represents the outcome change per SD increase in protein level; for proteins that decreased after torcetrapib treatment, beta represents the outcome change per SD decrease in protein level. For continuous outcomes, beta is reported as SD change in the outcome trait; for binary outcomes, beta is reported as the natural logarithm of the odds ratio.

##### Aptamer Selection

Protein QTL results from the ARIC and deCODE pQTL studies were identified for the differentially expressed proteins from ‘Supplemental Table II’ in Williams, Ganz and colleagues. Differential proteins were measured using 1129 aptamers from SomaLogic, which have been previously described.^<u>13,25</u>^ Aptamers for these proteins were matched to those from SomaScan version 4, which were used to identify pQTLs in ARIC and Icelanders. Aptamers were mapped using Uniprot IDs and Entrez gene symbols. When multiple aptamers were available for a given protein, the aptamer with the lowest pQTL p-value in ARIC was selected. When the aptamer was reported to bind more than one target protein, and each protein was targeted by individual aptamers on the SomaScan version 4 platform, the aptamers for each protein were retained. Protein QTLs were not available for seven protein targets from Williams et al.: HSD17B10, multitarget aptamers ‘PRKAA2 PRKAB2 PRKAG1‘, ‘CDK8 CCNC’ and ‘C4A C4B’ and the protein isoforms E2, E3 and E4 for APOE.

#### Assessment of instrumental variable assumptions

We tested the first instrumental variable assumption (relevance) by estimating the partial F-statistic, which tests the association of each pQTL with the protein. pQTLs with F-statistics of less than ten were dropped. We attempted to falsify the third instrumental variable assumption (exclusion) by investigating whether there was evidence of horizontal pleiotropy by removing variants with stronger associations with other proteins in the ARIC pQTL Study.

#### Outcome GWAS

We estimated the effects of each of the 95 proteins on 19 health outcomes using summary Mendelian randomization. We estimated the effects on the risk of adverse cardiovascular events, including coronary heart disease, ischemic stroke, heart failure, and peripheral arterial disease. We estimated the effects on cardiovascular disease risk factors, including HDL, LDL, triglycerides^<u>26</u>^, BMI^<u>27</u>^, HR^<u>28</u>^, systolic and diastolic blood pressure^<u>29</u>^, type-2 diabetes^<u>30</u>^, and a measure of eGFR^<u>31</u>^. We also investigated the effect of these proteins on three infection outcomes (infection, pneumonia, sepsis^<u>32</u>^), one autoimmune outcome (inflammatory bowel disease)^<u>33</u>^, and two cancers ( breast and prostate^<u>34</u>^). Please see Supplementary Table 3 for the GWAS used in this analysis. Summary data from each GWAS were downloaded and harmonized to the same effect allele and reference panel. We then used the TwoSampleMR package to extract the pQTLs from each outcome GWAS. Sample overlap: Very few samples overlap between the exposure and outcome GWAS, which is unlikely to materially affect the results.^<u>35</u>^

##### Instrument effects on transcription levels

To investigate tissue-specific effects of our genetic instruments, we examined whether variants used to instrument PDE5A serum protein levels also acted as expression quantitative trait loci (eQTLs) for PDE5A transcript levels. We queried eQTL associations across multiple tissues using data from the Genotype-Tissue Expression (GTEx) project version 8 (v8)^<u>36</u>^ and in whole blood from the eQTLGen Consortium.^<u>37</u>^

### Statistical analysis

#### Effect of proteins on the outcomes

Our primary analysis estimated the effects of each protein on each outcome using the fixed-effects inverse-variance weighted (IVW) estimator.^<u>38</u>^ This estimator assumes no horizontal pleiotropic effects but is one of the most precise summary data estimators.

#### Effect of the torcetrapib on each outcome

We used the Mendelian randomization estimates above to estimate the effects of the proteomic changes induced by torcetrapib by rescaling them to reflect the protein change caused by torcetrapib.

#### Reverse Mendelian randomization

We used reverse Mendelian randomization to estimate the effects of each of the 19 health outcomes on the proteome. We used the same parameters for clumping the variants as above and published GWAS for each disease.

#### Multiple testing

We used a Bonferroni-corrected significance level of 0.05 divided by the number of proteins and outcomes.

### Sensitivity analyses and additional analyses

We investigated the sensitivity of our results to the assumptions about the structure of pleiotropy by rerunning our analyses using the weighted median, weighted mode, and MR-PRESSO estimators, which relax assumptions around horizontal pleiotropy. We report whether each result was consistent across these estimators and the IVW analysis.

### Software, pre-registration, reporting

All analyses were conducted using R. We have reported this study using the STROBE-MR checklist.^<u>39</u>^

## Contributions

J.A.B., J.S.F. and N.M.D designed the study and drafted the manuscript. T.Y., C.M.S. and N.M.D. directed statistical analyses and provided methodological oversight. C.M.S., T.Y., B.M.P., and P.G. critically reviewed the manuscript.

## Data availability

We did not have access to any individual-level participant data for this study. The summary data used in the study can be accessed through the resources listed in Supplementary Data 3.

## Code availability

Code used to perform these analyses will be available on publication on GitHub: https://github.com/j-brody/drug-target-mr-analysis.

---

**Supplemental Figure 1:**
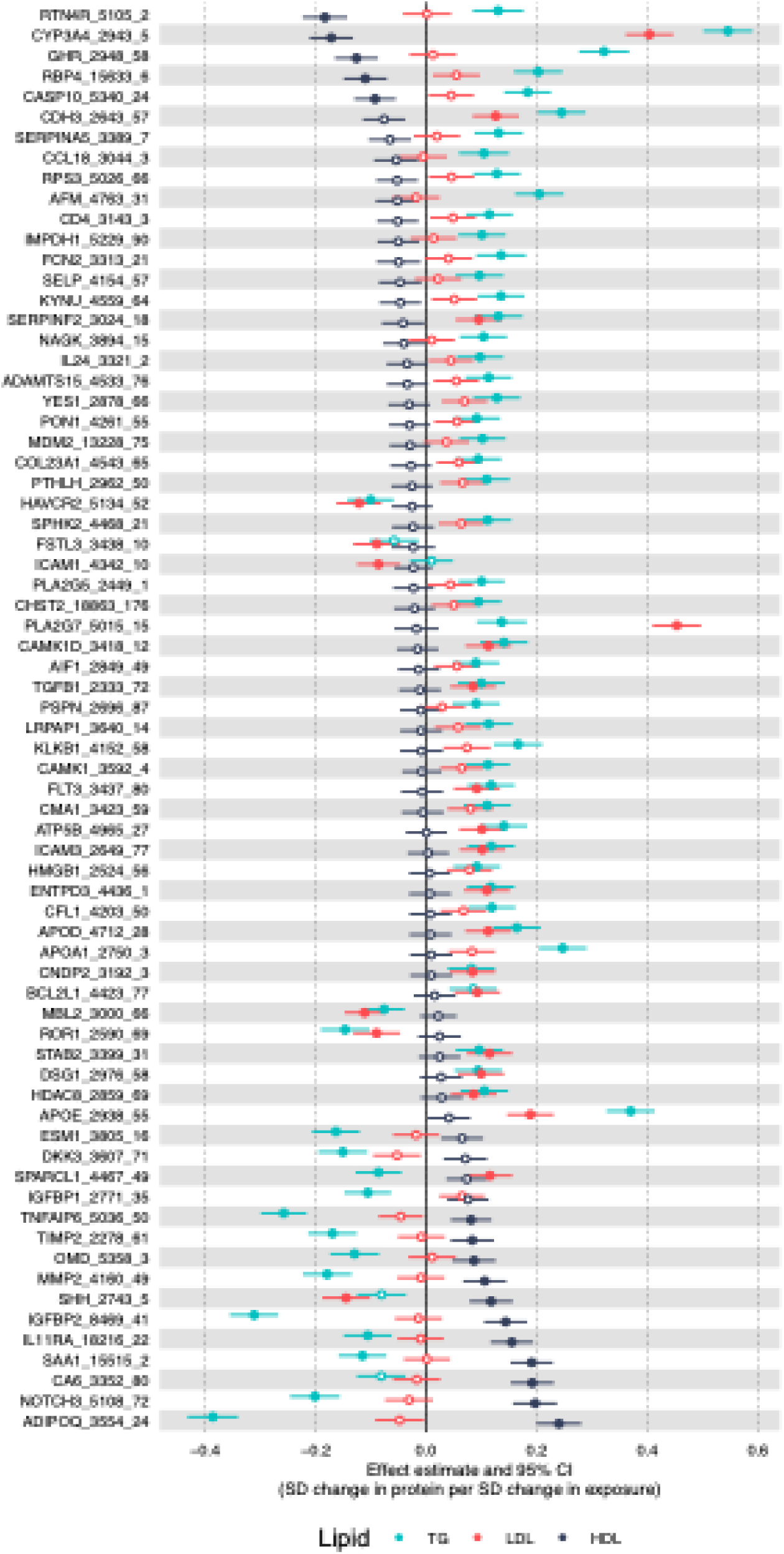
MR of lipid levels on proteins identified 70 proteins of the 200 differentially expressed proteins were causally impacted by lipid levels. Forest plot showing Mendelian randomization estimates for the causal effects of genetically predicted lipid levels (LDL cholesterol, HDL cholesterol, and triglycerides) on protein levels. Effect sizes represent the change in protein levels (in standard deviations) per standard deviation increase in each lipid fraction. Seventy proteins showed significant associations with at least one lipid fraction after Bonferroni correction for three lipids and 196 tested proteins. Proteins are ordered by ascending HDL cholesterol effect size. Points represent effect estimates with 95% confidence intervals: filled points indicate significant associations, and open points indicate non-significant associations.

## References

1. Kiriiri, G. K., Njogu, P. M. & Mwangi, A. N. Exploring different approaches to improve the success of drug discovery and development projects: a review. Futur J Pharm Sci 6, 27 (2020).

2. Wong, C. H., Siah, K. W. & Lo, A. W. Estimation of clinical trial success rates and related parameters. Biostatistics 20, 273–286 (2019).

3. Russo, L. et al. Genetic and other omics-based information in the most-cited recent clinical trials. Preprint at 10.1101/2024.10.21.24315878 (2024).

4. Minikel, E. V., Painter, J. L., Dong, C. C. & Nelson, M. R. Refining the impact of genetic evidence on clinical success. Nature 629, 624–629 (2024).

5. Davies, N. M., Holmes, M. V. & Davey Smith, G. Reading Mendelian randomisation studies: a guide, glossary, and checklist for clinicians. BMJ k601 (2018) doi:10.1136/bmj.k601.

6. Schmidt, A. F. et al. Genetic drug target validation using Mendelian randomisation. Nat Commun 11, 3255 (2020).

7. Pierce, B. L. & Burgess, S. Efficient Design for Mendelian Randomization Studies: Subsample and 2-Sample Instrumental Variable Estimators. American Journal of Epidemiology 178, 1177–1184 (2013).

8. Anderson, E. L. & Williams, D. M. Drug target Mendelian randomisation: are we really instrumenting drug use? Diabetologia 66, 1156–1158 (2023).

9. Toribio, M. et al. Assessing statin effects on cardiovascular pathways in HIV using a novel proteomics approach: Analysis of data from INTREPID, a randomized controlled trial. EBioMedicine 35, 58–66 (2018).

10. deFilippi, C., et al. Differential Plasma Protein Regulation and Statin Effects in Human Immunodeficiency Virus (HIV)-Infected and Non-HIV-Infected Patients Utilizing a Proteomics Approach. The Journal of Infectious Diseases 222, 929–939 (2020).

11. Kirsher, D. Y. et al. The Current Landscape of Plasma Proteomics: Technical Advances, Biological Insights, and Biomarker Discovery. Preprint at 10.1101/2025.02.14.638375 (2025).

12. Barter, P. J. et al. Effects of Torcetrapib in Patients at High Risk for Coronary Events. N Engl J Med 357, 2109–2122 (2007).

13. Williams, S. A. et al. Improving Assessment of Drug Safety Through Proteomics: Early Detection and Mechanistic Characterization of the Unforeseen Harmful Effects of Torcetrapib. Circulation 137, 999–1010 (2018).

14. Dalvie, D. et al. Pharmacokinetics, Metabolism, and Excretion of Torcetrapib, a Cholesteryl Ester Transfer Protein Inhibitor, in Humans. Drug Metabolism and Disposition 36, 2185–2198 (2008).

15. Fleming, T. R. & Powers, J. H. Biomarkers and surrogate endpoints in clinical trials. Stat Med 31, 2973–2984 (2012).

16. Trajanoska, K. et al. From target discovery to clinical drug development with human genetics. Nature 620, 737–745 (2023).

17. Schmidt, A. F. et al. Cholesteryl ester transfer protein (CETP) as a drug target for cardiovascular disease. Nat Commun 12, 5640 (2021).

18. Pietzner, M. et al. Synergistic insights into human health from aptamer- and antibody-based proteomic profiling. Nat Commun 12, 6822 (2021).

19. Katz, D. H. et al. Proteomic profiling platforms head to head: Leveraging genetics and clinical traits to compare aptamer- and antibody-based methods. Sci Adv 8, eabm5164 (2022).

20. Labrecque, J. A. & Swanson, S. A. Commentary: Mendelian randomization with multiple exposures: the importance of thinking about time. International Journal of Epidemiology 49, 1158–1162 (2020).

21. Walker, V. et al. Reading and conducting instrumental variable studies: guide, glossary, and checklist. BMJ e078093 (2024) doi:10.1136/bmj-2023-078093.

22. Zhang, J. et al. Plasma proteome analyses in individuals of European and African ancestry identify cis-pQTLs and models for proteome-wide association studies. Nat Genet 54, 593–602 (2022).

23. Hemani, G. et al. The MR-Base platform supports systematic causal inference across the human phenome. eLife 7, (2018).

24. Ferkingstad, E. et al. Large-scale integration of the plasma proteome with genetics and disease. Nat Genet 53, 1712–1721 (2021).

25. Gold, L. et al. Aptamer-based multiplexed proteomic technology for biomarker discovery. PLoS One 5, e15004 (2010).

26. Graham, S. E. et al. The power of genetic diversity in genome-wide association studies of lipids. Nature 600, 675–679 (2021).

27. Yengo, L. et al. Meta-analysis of genome-wide association studies for height and body mass index in ∼700000 individuals of European ancestry. Hum Mol Genet 27, 3641–3649 (2018).

28. van de Vegte, Y. J. et al. Genetic insights into resting heart rate and its role in cardiovascular disease. Nat Commun 14, 4646 (2023).

29. Evangelou, E. et al. Genetic analysis of over 1 million people identifies 535 new loci associated with blood pressure traits. Nat Genet 50, 1412–1425 (2018).

30. Suzuki, K. et al. Genetic drivers of heterogeneity in type 2 diabetes pathophysiology. Nature 627, 347–357 (2024).

31. Stanzick, K. J. et al. Discovery and prioritization of variants and genes for kidney function in >1.2 million individuals. Nat Commun 12, 4350 (2021).

32. Butler-Laporte, G. et al. Increasing serum iron levels and their role in the risk of infectious diseases: a Mendelian randomization approach. Int J Epidemiol 52, 1163–1174 (2023).

33. Liu, J. Z. et al. Association analyses identify 38 susceptibility loci for inflammatory bowel disease and highlight shared genetic risk across populations. Nat Genet 47, 979–986 (2015).

34. Sato, G. et al. Pan-cancer and cross-population genome-wide association studies dissect shared genetic backgrounds underlying carcinogenesis. Nat Commun 14, 3671 (2023).

35. Sadreev, I. I. et al. Navigating Sample Overlap, Winner’s Curse and Weak Instrument Bias in Mendelian Randomization Studies Using the UK Biobank. http://medrxiv.org/lookup/doi/10.1101/2021.06.28.21259622 (2021) doi:10.1101/2021.06.28.21259622.

36. The GTEx Consortium et al. The GTEx Consortium atlas of genetic regulatory effects across human tissues. Science 369, 1318–1330 (2020).

37. Võsa, U. et al. Large-scale cis- and trans-eQTL analyses identify thousands of genetic loci and polygenic scores that regulate blood gene expression. Nat Genet 53, 1300–1310 (2021).

38. Ye, T., Shao, J. & Kang, H. Debiased Inverse-Variance Weighted Estimator in Two-Sample Summary-Data Mendelian Randomization. Preprint at 10.48550/ARXIV.1911.09802 (2019).

39. Skrivankova, V. W. et al. Strengthening the reporting of observational studies in epidemiology using mendelian randomisation (STROBE-MR): explanation and elaboration. BMJ n2233 (2021) doi:10.1136/bmj.n2233.

